# Implementation of predictive risk stratification to reduce emergency admissions to hospital: experiences of general practitioners and practice managers

**DOI:** 10.1101/2021.01.25.21250442

**Authors:** Bridie Angela Evans, Jan Davies, Jeremy Dale, Hayley Hutchings, Mark Kingston, Alison Porter, Ian Russell, Victoria Williams, Helen Snooks

## Abstract

**Aim:** In a trial evaluating the introduction of a predictive risk stratification model (PRISM) into primary care, we reported statistically significant increases in emergency hospital admissions and use of other NHS services without evidence of benefits to patients or the NHS. The aim of this study was to explore the views and experiences of general practitioners (GPs) and practice managers on incorporating PRISM into routine practice.

**Methods:** We interviewed 22 GPs and practice managers in 18 participating practices at two timepoints: 3-6 months after PRISM was available in their practice; and at study end, up to 18 months later. We recorded and transcribed interviews and analysed data thematically using Normalisation Process Theory.

**Results:** Respondents reported that the decision to use PRISM was based mainly on fulfilling reporting requirements for Quality and Outcome Framework (QOF) incentives. Most applied it to a very small number of patients for a short period. Using PRISM entailed technical tasks, information sharing within practice meetings and changes to patient care. These were diverse and generally small scale. Use was inhibited by PRISM not being integrated with practice systems. Respondents’ evaluation of PRISM was mixed: most doubted it had any large scale impact, but many cited examples of impact on individual patient care. They reported increased awareness of patients in high risk groups.

**Conclusions:** Qualitative results suggest mixed views of predictive risk stratification in primary care and raised awareness of highest-risk patient groups, potentially affecting unplanned hospital attendance and admissions. To inform future policy, decision-makers need more information about implementation and effects of emergency admissions risk stratification tools in primary and community settings.

**Trial registration:** Controlled Clinical Trials no. ISRCTN55538212.

## Background

Predicting risk of emergency hospital admission using computer software is widely advocated to support proactive management of the care of vulnerable patients and manage demand on healthcare services [1-4]. In 2012–2013, there were 5.3 million emergency admissions to hospitals in England costing approximately £12.5 billion [5]. Around half of these admissions arise from 5% of the population—typically older people with comorbidities [6]. Patients with chronic conditions are at increased risk of flare-ups and unplanned hospital attendance and admission, yet an estimated one in five emergency admissions is avoidable [7, 8]. In addition to being costly, an emergency admission to hospital is disruptive and carries risks to patients’ clinical and psychological health. Health policies recommend proactive management of conditions amenable to community prevention or care [9, 10].

Predictive risk stratification identifies patients at high risk of emergency admission to hospital, so that clinicians can deliver proactive management [8, 10, 11, 12, 13, 14]. Individual risk scores are estimated with predictors relating to past use of healthcare, diagnoses and medications. When combined with a software platform, these models underpin clinical risk prediction tools which support case finding and resource allocation and can allow targeting of different types of care and services to people at different levels of risk [15]. Such tools are generally more accurate and consistent than clinical opinion in identifying patients at risk of unplanned hospital admission [16]. The rationale for implementing predictive risk stratification tools is that targeted management of patients can reduce emergency admission, improve patient outcomes and experience and provide better value for money [14, 17]. However, to date there is little evidence that case management is effective in reducing secondary care use or costs, nor about the role of predictive risk stratification in case management [18]. Evidence regarding implementation and effects on quality and safety of care is also weak [12, 18]. Predictive risk stratification in combination with case management is widely promoted in policy internationally and across the UK [11, 12, 13, 19, 20, 21]. Recent incentive schemes have focused on patients identified at the highest level of risk [22, 23].

We evaluated the implementation and effect of one emergency admissions risk stratification tool (PRISM) across 32 GP practices in Wales (NIHR Health Services Delivery and Research programme: project no. 09/1801/1054) [24]. The PRISMATIC trial, using a randomised stepped wedge design [10, 25], found that, contrary to expectations, the introduction of predictive risk stratification increased emergency admissions to hospital; full results are reported elsewhere [26]. A qualitative component of the study aimed to describe the processes of change associated with PRISM: how it was understood, communicated, adopted, and used by practitioners, managers, local commissioners and policy makers.

Implementing new health care technologies such as risk prediction tools can be slow and difficult [27, 28]. Normalisation Process Theory (NPT) [29] is increasingly used as a conceptual framework to examine and explain this [30]. NPT considers implementation as a process which entails sustained work by those responsible and suggests four constructs that help us understand how innovation happens in routine practice: how people understand the innovation and its purpose (coherence); what decisions are taken to use it, based on perceived advantages (cognitive participation); what people do to bring the innovation into everyday use (collective action); how an innovation is reviewed, modified or abandoned (reflexive monitoring) [31].

We have previously reported on coherence, the first of these constructs, explored in focus groups with GPs and other practice staff conducted before PRISM was introduced [32]. They welcomed the opportunity to use the tool, believing it could help them manage burgeoning workloads and increase the sustainability of general practice. Some dubbed it a ‘golden goose’ for its potential both to benefit patients and manage demand on health services.

Here we use the other three constructs of NPT (cognitive participation, collective action, reflexive monitoring) to shape our analysis of the reported experiences and reflections of GPs and Practice Managers after they were given access to the PRISM tool and how they used it in their daily practice.

### Aim

To explore views and experiences of general practitioners and practice managers who used the PRISM risk stratification tool.

### Setting

The PRISMATIC stepped wedge trial took place in 32 practices in one largely urban area in South Wales. Each practice nominated a GP lead responsible for coordinating use of PRISM and participation in PRISMATIC, including engagement with other clinical and practice staff. We invited GPs to a practice-based training session about using PRISM to identify patients at risk of emergency admission. We provided a user-friendly handbook and access to clinical support through two locally appointed ‘GP champions’ plus technical support via email or telephone to the Primary Care Service Desk at NHS Wales Informatics Service. We advised individual practices that they could choose how to use the tool within their practice.

During the study period, the Welsh Government introduced a financial incentive, through the QOF, to encourage GPs to use emergency admission risk stratification tools to support hospital avoidance [23](Table 1 and Appendix 1). The Health Board covering the participating general practices encouraged them to use the PRISM tool to undertake this work. The phased rollout according to the stepped wedge design meant practices had access to PRISM for different lengths of time during this period.

**Table 1:** summary of QOF tasks to support hospital avoidance

|  |
| --- |
| • Produce a list of 5% of patients in the practice who are predicted to be at significant risk of unscheduled admission<br>• Review 10% of patients on this list (maximum 0.5% of practice list) and prepare an active management plan<br>• Hold at least four meetings a year to review care for identified patients |

## Methods

We purposively sampled half the participating practices to cover a range of practice sizes and geographic spread. We interviewed the lead GP in each at two timepoints: 1) between three and six months after PRISM was activated in their practice; 2) at the end of the intervention phase, about 18 months after it was available in the first practices. This enabled us to understand how the tool was introduced and used and to describe changes over time. In a few practices, the Practice Manager contributed to these interviews.

Two experienced researchers from the study team (BAE and MRK) conducted all interviews, held at general practices and lasting between 30 and 90 minutes. We provided information about the aim of the study. With participants’ consent, we recorded and transcribed interviews. We made field notes after each interview. See Appendix 2 for interview schedules.

### Analysis

We analysed interview transcripts thematically, informed by NPT as our underlying theoretical framework. Thematic analysis is a systematic and transparent method that generates themes from the explicit and implicit ideas in the original accounts of participants [33]. Team members (researchers BAE, MRK, AP, VW and service user SW) read the transcripts and developed a coding framework. BAE led the analysis with the others independently supporting key stages of coding, generating themes and interpretation, thus encouraging a critical stance to test and confirm findings [34, 35, 36].

### Reporting

We selected quotations to be illustrative and typical of respondents’ comments unless otherwise stated. Where a respondent emphasised a word or phrase, that emphasis is indicated by bold type. Quotations are identified by respondent role (GP for general practitioner, PM for Practice Manager), practice unique identifier, time point (interview 1 or interview 2). For example: ‘GP01 interview1’ would identify a quotation from the first interview with the GP from practice number 1. ‘

### Service user involvement

We report service user involvement according to GRIPP guidance [37]. We recruited two service users (SW, JanD) who acted throughout the study as collaborators in our research partnership [38, 39, 40]. As members of the Research Management Group, they attended the quarterly meetings responsible for strategic and operational decisions about the study. They contributed as equal team members in all meetings to ensure we considered patients’ perspectives at all stages of the study. SW was also involved in analysing the qualitative data.

We recruited SW and JanD through SUCCESS (Service Users with Chronic Conditions Encouraging Sensible Solutions), a group of patients and carers engaged in research linked to the chronic conditions management policy in Wales (http://www.invo.org.uk/posttypeconference/an-alternative-success-model/). They kept PRISMATIC linked to SUCCESS by reporting regularly to SUCCESS and seeking feedback to inform their contributions [41].

We recruited two more service users to the Trial Steering Committee through Involving People (http://www.wales.nhs.uk/sites3/page.cfm?orgid=1023&pid=59261) to ensure their independence. We followed best practice by ensuring all users received honoraria, expenses, training and support, a named contact, information and networking opportunities [42].

### Ethics

We obtained full ethical approval for the main protocol and all subsequent amendments from the Multi-Centre Research Ethics Committee for Wales (reference 10/MRE09/25).

## Results

We conducted 22 interviews at timepoint 1 and 19 interviews at timepoint 2 (Table 2). Most respondents were GPs. One GP left between the first and second interviews while two Practice Managers were not available at the second interview.

**Table 2:**
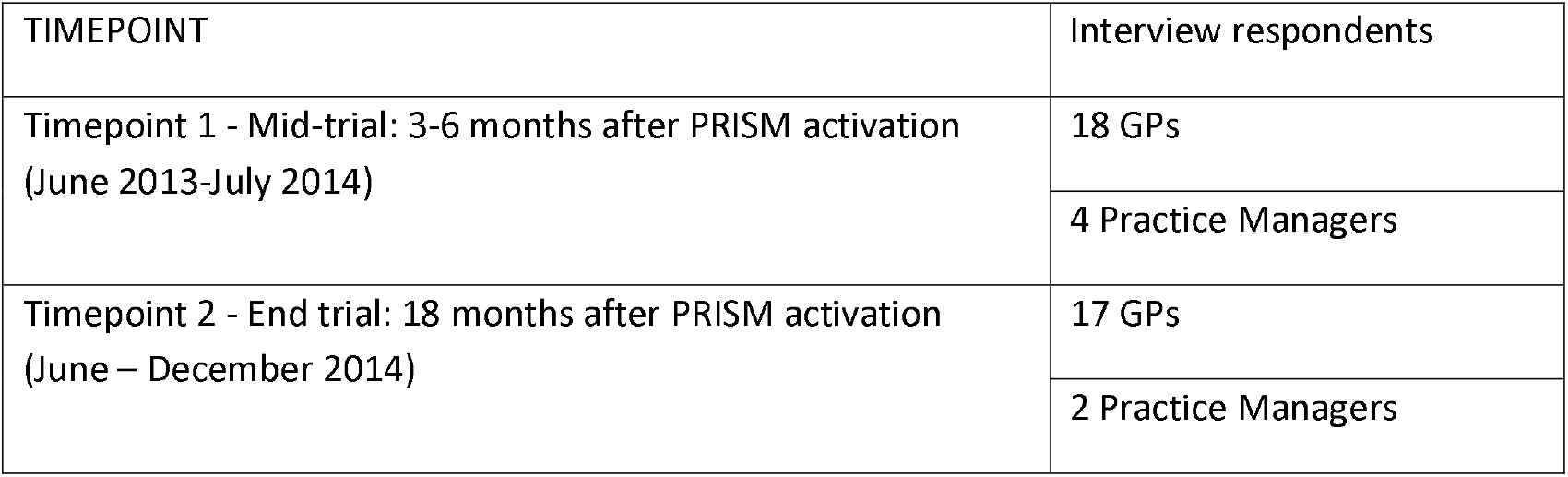
Numbers and roles of respondents at data collection timepoints

| TIMEPOINT | Interview respondents |
| --- | --- |
| Timepoint 1 - Mid-trial: 3-6 months after PRISM activation<br>(June 2013-July 2014) | 18 GPs |
|  | 4 Practice Managers |
| Timepoint 2 - End trial: 18 months after PRISM activation<br>(June – December 2014) | 17 GPs |
|  | 2 Practice Managers |

We present results showing how staff reported using PRISM tool and their reflections on their practice and patient care. We cover results for the three remaining constructs of Normalisation Process Theory – cognitive participation, collective action and reflexive monitoring - having covered coherence elsewhere [32].

### Cognitive participation: deciding to use PRISM

Respondents described how they made decisions about using PRISM, based on its perceived advantages. There was a consistent message that they used it mostly to identify patients at high risk of emergency admissions to fulfil QOF requirements.

> *‘it was fantastic because we were able to pick out the patients that the local health board had highlighted for the QOF thing’ (GP11 interview2)*

A few practices reviewed and refreshed their PRISM list throughout the trial period. Respondents said the majority of patients were known and considered high risk, but some names were unexpected or unfamiliar. During the second interview at the end of the study, only two practices reported that they were still using PRISM after they completed and submitted their QOF reports.

A few GPs, who had earlier access to PRISM and longer to use the tool, reported using PRISM to support patient care in ways outside the QOF requirements. They interrogated the PRISM data to understand the reason for a high score or to review specific patient groups such as COPD patients.

> *‘looking at patients that’ve been deemed high risk, but not necessarily the highest risk, people that we might be able to do something about. Just exploring the data and seeing if there is anything that we could do to be more proactive*.*’ (GP13 interview2)*

### Collective action: what people did to bring PRISM into use

Bringing PRISM into use involved three processes: using the technology itself; sharing the information it generated with relevant clinical teams; and, in the light of this information, taking action with patients. Typically, the lead GP or Practice Manager generated a list of patients within the top stratum of risk and saved this in an electronic spreadsheet or printed it. A few practices also created a screen popup on the record of high risk patients, which alerted all staff to tailor care or make early appointments when these patients phoned reception.

> *‘that flags up – that’s like a warning sign that maybe we should take them a bit more seriously’ (GP06 interview2)*

PRISM was not without technical challenges: slow broadband speeds; system crashes; passwords locking or not working. Some respondents also complained that PRISM was not integrated with practice-based clinical information, inhibiting routine management of patients’ data:

> *‘what I wanted was to download my 53 patients…the information that would allow me to work out why they’re on that list…and it really disappointed me that a lot of that I had to do manually’ (GP15 interview1)*

Respondents described sharing the list of individual patients’ risk scores at routine practice meetings for discussion when time allowed, or dividing it among partners who worked individually, if close to the QOF deadline.

More than half the respondents said the work of bringing PRISM into use was inhibited by the many demands on GPs’ time, shortage of GPs due to illness, retirement or maternity leave; and a couple of practices were planning to close or merge.

GPs reported a range of actions to care of high-risk patients after using PRISM. These were generally small amendments to supplement existing treatment and support for individual patients, or extra reviews (some through house calls) to fine-tune existing treatment.

> *‘There will have been people who were reviewed or assessed, who otherwise wouldn’t have been’ (GP14 interview2)*

Some reviews explicitly focused on how best to manage crises which might lead to emergency admission, with one GP reporting the conversation with a patient during a face to face consultation:

> *‘“Look – you’ve been admitted on a number of occasions. Obviously the chances of you being admitted are quite high, why don’t we do something a bit different? We will try and alter your medication to maybe control your condition a bit better. We are here during the day, so use us rather than dial 999, and we can get somebody to see you*.*” ‘(GP11 interview2)*

Other effects on patient care were reported. In one practice the nurse talked to high risk patients about identifying infections, weight management and spotting early warning signs. In another, emergency drug packs were made up for identified patients to use at weekends and bank holidays if they felt their heath was deteriorating. Some high risk patients were referred to outpatient clinics, nursing teams or other care agencies for non-medical needs. Some respondents said they were concerned that practice staff did not have the capacity or skills to take on more patients. All respondents reported difficulty changing from a reactive to a proactive approach to care when the daily routine in general practice was so busy and GPs said they felt overworked.

### Reflexive monitoring: reviewing PRISM

Despite early optimism that PRISM could support more proactive care, GPs generally judged it unlikely that PRISM had any effect on emergency admissions and ED attendances. There was a widespread feeling that admissions initiated by GPs were already low and could not be reduced further. They did not believe additional contact with patients identified as being at high risk could change this, though they were very aware of media coverage about emergency use of services and said they were concerned to use resources appropriately and carefully.

> *‘There are odd occasions where you were able to proactively help somebody or put a plan in place to stop them being admitted to hospital. I think it’s a fallacy to think that you could reduce emergency admissions from primary care because the primary care admissions are so small*…*one case a week, if that*.*’ (GP02 interview2)*

A minority of respondents could identify instances where an emergency admission may have been avoided and two GPs who targeted patients with frequent ED attendance reported that those patients’ use of 999 services had fallen. In contrast, one GP suggested the PRISM reviewing may have been associated with increased hospital admissions.

> *‘We did bring in a certain number of people and do care plans with them and then we ended up admitting them because we’d seen them and they looked unwell*.*’ (GP31 interview2)*

Reflecting on the effect of PRISM, the majority of respondents described how it changed their awareness of high risk patients, especially when unexpected patients appeared in the top stratum.

> *‘We’ve probably had had more talks together, as a group, as to how we have contacted the ambulance service and A & E*… *And we’ve talked about these patients more I think*.*’ (GP18 interview2)*

GPs said the combination of PRISM and the QOF incentive increased their contact with some patients in this group, explaining that they did this to reassure themselves that these patients’ care was optimum.

> *‘It [a high risk score] does have an effect of making you sit up and think ‘heck, what’s he doing up?’ (GP05 interview2)*

Most believed that the increased GP-patient interaction was probably beneficial, regardless of any treatment delivered, because patients appeared to appreciate the extra attention and, where appropriate, tailored self management advice. GPs did not generally tell patients about PRISM or share the risk score, careful not to alarm and precipitate self-referrals.

Some respondents felt the QOF focus on highest-risk patients was misplaced. They believed these few patients was already well known to the practice, but that medium-risk patients had the most to gain from close attention and proactive care, if resources were available.

> *‘Those in the middle bracket were the kind of patients that you were possibly able to help more than those*…*in the higher echelons that were already having all the input that was available. But it was the ones in the next cohort down which might have been more useful because you can actually put in things that will stop them from going up the pyramid*.*’ (GP02 interview2)*

Respondents reflected that the QOF payments encouraged short term use of PRISM, in the absence of extra resources to support changing practice in the long term. The network of community health and social services needed to support proactive care were generally not available for the high risk patients they reviewed.

> *‘We discussed those patients at various meetings, we made plans about how to minimise admissions, access Out of Hours, Casualty, but it’s not in this year’s QOF. We’re far too busy to carry on with something I didn’t really see derive any particular benefit for anybody, because we were aware of those patients, and it’s been many years now that there’s been an emphasis not to admit*… *So, it was more a question of a useful tool to achieve points, more than anything else*.*’ (GP10 interview2)*

## Discussion

### Summary of findings

Our study identified a range of often contradictory views about the use and usefulness of PRISM within general practices. GPs and Practice Managers reported that the decision to use PRISM was based mainly on fulfilling QOF requirements. For most, it was applied to a very small number of patients with high risk of emergency admissions for a short period because that was a contractual requirement. Only a minority used PRISM in other ways, such as identifying patients at the medium risk level. Despite considering that they knew their high-risk patients well and they would not benefit greatly from more resources, GPs said their awareness of these individuals was heightened by knowing the PRISM scores. They also provided more care to this group.

The work of bringing PRISM into practice had technical aspects, inhibited by the fact that it was not integrated with practice systems. It also entailed information sharing, generally done in practice meetings. It seemed the QOF incentive to use the tool temporarily overcame systemic barriers such as other demands on GPs time, shortage of GPs, software and technical problems. Changes to care of high-risk patients as a result of using PRISM were diverse and generally small scale, such as extra visits, care plan reviews, medication amendments, tailored self-care advice and referrals to other services. Respondents’ evaluation of PRISM was mixed: there were doubts about it having any large scale effect, but many cited effects on individual patients. Some concerns were expressed about way QOF promoted use of PRISM, particularly the focus on highest risk patients who may have been least suitable for proactive management, and the short term nature of the implementation.

### Strengths and limitations

We interviewed GPs and Practice Managers with a wide range of experiences across 18 diverse practices in rural, urban, prosperous and deprived communities. Collecting data at two time points allowed us to recognise changes in attitudes and expectations. However, we mainly talked to one individual per practice and relied on their reports of views and activities of other practice staff.

These practices volunteered to take part in the PRISMATIC trial, committing themselves to using the PRISM tool for a fixed time and receiving a small payment to acknowledge the additional demands on time of practice staff associated with participation in the study. Their views may not be typical of practices which did not take part in the study.

We examined participation, action and reflections among GPs who used PRISM in the front line. We do not know the views of local commissioners and managers responsible for services for patients at risk of emergency admission. Nor do we know why commissioners and managers required practices to use PRISM to achieve the QOF requirements.

### Implications for policy, practice and research

Software in general practices that predicts risk of emergency hospital admission for every registered patient has been widely promoted as a means of targeting preventive services to people at high risk to reduce crises that result in emergency admissions [10, 22].However main PRISMATIC findings showed that introduction of PRISM resulted in a statistically significant increase in emergency hospital admissions and use of other NHS services without evidence of benefits to patients or the NHS [26]. One interpretation is that the increases arose from changed awareness and behaviour, among GPs and other practice staff, particularly when individuals they hadn’t expected arrived in the top stratum, making them more cautious in their clinical practice. This may have resulted in over-investigation, over-diagnosis and over-treatment which may in turn lead to potentially serious side effects including emergency admissions. It is also possible that patients (and their carers) who received extra contact became more aware of their poor health and sought emergency care when they previously would not have done. Health anxiety is one factor in a patient’s decision to seek emergency care [43]. Additionally, it may be that GPs had less time for other patients. Because of their extra contact with those at highest risk, health needs of other patients may not have been promptly seen and they could have sought emergency hospital attention [44]. As our study was highly powered, moderate behaviour changes at individual surgeries contributed to significant increases in NHS activity across the 32 participating practices. These sick patients are also more likely to be admitted to hospital in an emergency and remain for some time [45, 46].

Predictive risk stratification software is now widely available in primary care across the UK [47], though existing research evidence does not support its use as a means of reducing emergency admissions to hospital [18, 25, 26]. PRISMATIC findings illustrate the unpredictable consequences of introducing service innovations into NHS practice. Our qualitative data do not entirely explain our quantitative findings of a rise in NHS activity when the risk prediction tool arrived in general practices [26]. But they do offer insight into the changed perceptions and behaviours of general practice staff and help to understand the unexpected trial results. They also highlight how complex are the processes of adopting and using innovation in the real world and how apparently helpful incentive schemes may distort outcomes. Embedding this qualitative work within the PRISMATIC trial responded to calls for thorough understanding of how new services are adopted and used, since organisational culture affects implementation [48, 49]. Our findings highlight the need for further research into behaviours and attitudes within general practice to inform use of emergency admissions risk stratification tools as they are available [47].

## Conclusions

Emergency admission risk stratification tools are widely advocated to reduce emergency hospital admissions and are available in primary and community care across much of the UK. However, there is a lack of evidence to support the view that they enable proactive care and improve patient outcomes. We found varied views and experiences among GPs and Practice Managers about use of the PRISM tool, which was short term and driven by external factors rather than embedded in new ways of working. Raised awareness of patient risk and focusing attention on the small numbers of patients who are at greatest risk may explain quantitative trial findings of increased emergency hospital admissions and use of other NHS services. Decision-makers need more information about the implementation and effects of such emergency admissions risk stratification tools in primary and community settings to inform future policy on their use and negative effects on patients and the NHS.

## Data Availability

All data generated or analysed during this study are included in this published article.

## List of abbreviations

NHS: National Health Service
QOF: Quality and Outcomes Framework
UK: United Kingdom
NIHR: National Institute for Health Research
PRISMATIC: Predictive RIsk Stratification Model: A Trial In primary Care
PRISM: Predictive Risk Stratification Model
GP: General Practitioner
NPT: Normalisation Process Theory
SUCCESS: Service Users with Chronic Conditions Encouraging Sensible Solutions
ED: Emergency Department

## Declarations

### Ethics approval and consent to participate

We gained ethical approval from the Multi-Centre Research Ethics Committee for Wales (reference 10/MRE09/25). All respondents gave informed consent in writing to participate in this study.

### Consent for publication

No applicable

### Availability of data and materials

All data generated or analysed during this study are included in this published article.

## Authors’ contributions

BAE drafted the manuscript with editorial input from all authors – JanD, JD, HH, MK, AP, IR, VW, HS. BAE led qualitative analysis with JanD, MK, AP and VW. The research idea was conceived and developed by HH, IR, HS. All authors read and approved the final manuscript.

## Acknowledgements

We thank all the general practices who took part in PRISMATIC and the GPs and Practice Managers who gave their time to be interviewed by team members. We also acknowledge Shirley Whitman, one of two service users involved in undertaking PRISMATIC, who was unable to contribute to this paper.

