## Appendix 1 for "Implementation of predictive risk stratification to reduce emergency admissions to hospital: experiences of general practitioners and practice managers"

**Appendix 1: Quality and Productivity Indicators within Quality and Outcomes Framework (QOF) guidance for General Medical Services contract Wales 2013/14**

**Key tasks for a General Practice**

QP100W The practice produces a list of 5% of patients in the practice who are predicted to be at significant risk of unscheduled admission to secondary care or community based alternatives (10 points)

QP101W The practice identifies a minimum of 10% (with a maximum of 0.5% of the practice list) of those patients from the list produced in indicator QP15 who would most benefit from review and ensures there is an active management plan (see template attached) is in place for each patient. The active management plan must identify the lead clinician or care coordinator and an appropriate review date. The frequency of each patient's review should be determined in light of their clinical and care needs. The practice will be responsible for ensuring that an appropriate system is in place for monitoring and review of the patients identified. (10 points)

QP102W The practice has at least four meetings during the year to review the delivery of care for the patients identified in QP16. These meetings should be open to all relevant professionals engaged in the delivery of care to this group. The meetings should be used to review the clarity of care plans and the effectiveness of service delivery. Patient and carer feedback should play a key role in informing this assessment. Participants should seek to identify opportunities to improve systems of care and to enact changes where possible. (22.5 points)
