## Appendix 2 for "Implementation of predictive risk stratification to reduce emergency admissions to hospital: experiences of general practitioners and practice managers"

**Prism 3 month interview schedule**

To be administered to 50% of participating practices, between three and six months after they are able to start using Prism. **All questions in bold will be included in the interview schedule to be conducted nine months after practices can use Prism.**

Objective: **Describe processes of change associated with Prism: how it is understood, communicated, adopted, and used by practitioners**, managers, local commissioners and policy makers.

1. Your practice received training to use Prism about three months ago. What happened next?
   - *Used immediately / delayed start*
   - *What decisions were made with colleagues – at PRISM training or after?*
   - *Who used it initially?*
   - *Not used*
     - *How easy / how difficult was it to start using Prism?*
2. What influence did the training have on your planning for and use of Prism?
   - Views on training session
   - Views on handbook
3. **Over the past three months, how has your practice been using Prism?**
   - ***How frequently***
   - ***How long do you usually spend each time you review Prism data***
   - ***Who has been involved***
     - ***Single user***
     - ***Routine practice meetings***
     - ***Special meetings to discuss Prism***
       - ***Who with – MDT, GPs, community network staff***
     - ***Informal meetings***
     - ***Clarify how long other people spend reviewing Prism data***
   - ***How do you use the information provided by Prism***
   - ***Do you review individual or grouped data***
   - ***Do you inform the patient that you are using Prism***
4. **Please think back to the most recent occasion you used Prism. Can you talk me through what you did?**
   - ***Used at screen / reviewed printed lists***
   - ***Alone / in meetings***
   - ***Which groups of patients (risk levels, conditions, other categories)***
   - ***What actions have you taken as a result of the information provided by Prism***
     - ***GP consultation in person***
     - ***GP consultation by telephone***
     - ***GP consultation by home visit***
     - ***Practice nurse appointment***
     - ***Clinic in GP practice (e.g. diabetes, asthma)***
     - ***Prepare Active Management Plan***
     - ***Communication with / refer to Community Resource Team***
     - ***Communication with / refer to district nurse***
     - ***Communication with / refer to health visitor***
     - ***Refer to community nurse/case manager (e.g. respiratory, mental health, chronic conditions)***
     - ***Communication with / refer to mental health services (identify which)***
     - ***Communication with / refer to therapist (e.g. physiotherapist, Occupational Therapist, speech therapist, rehabilitation)- identify which***
     - ***Communication with / refer to Social Services***
     - ***Refer to alternative medicine provider***
     - ***Refer to Voluntary Sector service (say what)***
     - ***Other………….(please give details)***
   - ***Have you needed to identify additional resources when taking action as a result of information provided by Prism? Probe what.***
5. **How has the way you use Prism changed during the past three months?**
   - ***Used by different people***
   - ***Started then stopped using Prism***
   - ***Only recently started using Prism***
   - ***Change in patients being reviewed through Prism***
   - ***Changed frequency of use – how?***
6. **Is there anything that has limited your use of Prism?**
   - ***Concerns about what it’s for/overall purpose***
   - ***Practical/technical difficulties with using it***
     - ***Issues with web access/password logins***
   - ***Concerns about accuracy – frequency of update, measures feeding into it***
   - ***Concerns about data handling and sharing***
   - ***Demands on time to access and understand***
   - ***Use as performance monitoring tool – compliance, liability***
   - ***Does not provide new information***
   - ***Unable to respond to identified needs***
7. **How do you feel about using Prism at the moment?**
   - ***Do you have a clear understanding of what the tool is for?***
   - ***Does it give you new/useful information***
   - ***Value for money?***
   - ***What has influenced your/your colleagues’ use of Prism?***
   - ***How does your experience match your expectations?***
   - ***Impact of QOF in deciding how you use Prism***
8. **What difference has Prism made to the way you work and patient care in the past three months?**
   - ***Why?***
   - ***Individual changes or organisational changes?***
   - ***Extent of your role in communicating / using Prism in the practice***
   - ***Do you think this is making a difference to emergency admissions?***
9. **How do you expect to use Prism over the next six months?**

- ***Don’t expect to use***
- ***Expect to use differently from first three months***
- ***Focus for discussion***
- ***New approach to dealing with patients***
- ***Expected strengths / weaknesses***
- ***Extent of your role in communicating / using Prism in the practice***
- ***Impact of QOF***

10. **Thank you for telling me what you think about Prism, If I talked to your colleagues, would they share your views or do they feel differently?**
    - ***Their experience of using it***
    - ***Their experience of impact on their work***
    - ***Their expectations for next six months***
    - ***Other GPs, practice nurse, practice manager etc.***

**Prism 9 month interview schedule**

To be administered to 50% of participating practices, at the end of the study period when they are able to use Prism. All practices will have already been interviewed about Prism three to six months after it was available in their practice.

Objective: **Describe processes of change associated with Prism: how it is understood, communicated, adopted, and used by practitioners**, managers, local commissioners and policy makers.

**If interviewing a different person from first interview respondent....**

**Last time we visited your practice, we talked to Dr .... Could you explain why this interview is taking place with you?**

- Change of staff (doctors left and arrived)
- Change of person leading Prism in the practice and why
- When did this change take place
- Any other staff changes in past six months?

11. **A. Last time we talked to you/your practice, you said you mainly used Prism for .....[check back to 3 month interview content]**

**Over the past six months, how has your practice been using Prism?**

- ***How frequently***
- ***How long do you usually spend each time you review Prism data***
- ***Who has been involved***
  - ***Single user***
  - ***Routine practice meetings***
  - ***Special meetings to discuss Prism***
    - ***Who with – MDT, GPs, community network staff***
  - ***Informal meetings***
  - ***Clarify how long other people spend reviewing Prism data***
- ***How do you use the information provided by Prism***
- ***Do you review individual or grouped data***
- ***Do you inform the patient that you are using Prism***
- ***Have you discussed individual Emergency Admissions risk scores with patients?***

12. **What has influenced the use of Prism in your practice in the past six months?**

- **Received guidance from**
  - **ABMU**
  - **Locality teams**
  - **Deb Burge-Jones**
- **Change of QOF/QPI requirements to identify high risk patients**
- **Identifying what works best in this practice**
- **Anything which has supported your use of Prism?**
- **Time of year (winter vs summer)**
- **Other influences on how you use Prism?**

13. **How has the way you use Prism changed during the past six months?**
    - ***Used by different people***
    - ***Started then stopped using Prism***
    - ***Only recently started using Prism***
    - ***Change in patients being reviewed through Prism***
    - ***Changed frequency of use – how?***
14. **Please think back to the most recent occasion you used Prism. Can you talk me through what you did?**
    - ***Used at screen / reviewed printed lists***
    - ***Alone / in meetings***
    - ***Which groups of patients (risk levels, conditions, other categories)***
    - ***Used in individual patient consultation***
    - ***What actions have you taken as a result of the information provided by Prism***
      - ***GP consultation in person***
      - ***GP consultation by telephone***
      - ***GP consultation by home visit***
      - ***Practice nurse appointment***
      - ***Clinic in GP practice (e.g. diabetes, asthma)***
      - ***Prepare Active Management Plan***
      - ***Communication with / refer to Community Resource Team***
      - ***Communication with / refer to district nurse***
      - ***Communication with / refer to health visitor***
      - ***Refer to community nurse/case manager (e.g. respiratory, mental health, chronic conditions)***
      - ***Refer to secondary care***
      - ***Communication with / refer to mental health services (identify which)***
      - ***Communication with / refer to therapist (e.g. physiotherapist, Occupational Therapist, speech therapist, rehabilitation)- identify which***
      - ***Communication with / refer to Social Services***
      - ***Refer to alternative medicine provider***
      - ***Refer to Voluntary Sector service (say what)***
      - ***Other………….(please give details)***
    - ***Have you needed to identify additional resources when taking action as a result of information provided by Prism? Probe what.***
    - ***Is this action typical of your normal use – what else have you done?***
15. **Is there anything that has limited your use of Prism?**
    - ***Concerns about what it’s for/overall purpose***
    - ***Practical/technical difficulties with using it***
      - ***Issues with web access/password logins***
      - ***Lack of integration with practice systems***
    - ***Concerns about accuracy – frequency of update, measures feeding into it***
    - ***Concerns about data handling and sharing***
    - ***Demands on time to access and understand***
    - ***Use as performance monitoring tool – compliance, liability***
    - ***Does not provide new information***
    - ***Unable to respond to identified needs***

***Last time we talked to you, you said ... limited your use of Prism. How has that changed?***

16. **How do you feel about using Prism at the moment?**
    - ***Do you have a clear understanding of what the tool is for?***
    - ***Does it give you new/useful information***
    - ***Value for money?***
    - ***What has influenced your/your colleagues’ use of Prism?***
    - ***How does your experience match your expectations?***
    - ***Impact of QOF in deciding how you use Prism***
17. **What difference has Prism made to the way you work and patient care in the past six months?**
    - **Compared to three months before?**
    - **Compared to pre-Prism?**
    - ***Why?***
    - ***Individual changes or organisational changes?***
    - ***Extent of your role in communicating / using Prism in the practice***
    - ***Do you think this is making a difference to emergency admissions?***
    - ***Do you think this is making a difference to any other outcomes?***

***When we talked to you six months ago, you said Prism made .... difference. Why do you think this has changed? (if there is a change)***

18. **How do you expect to use Prism over the next six months?**

- ***Don’t expect to use***
- ***Expect to use differently from past six months***
- ***Focus for discussion***
- ***New approach to dealing with patients***
- ***Expected strengths / weaknesses***
- ***Extent of your role in communicating / using Prism in the practice***
- ***Impact of QOF***
- ***Becoming a routine part of practice?***

19. **Thank you for telling me again what you think about Prism. If I talked to your colleagues, would they share your views or do they feel differently?**
    - ***Their experience of using it***
    - ***Their experience of impact on their work***
    - ***Their expectations for next six months***
    - ***Other GPs, practice nurse, practice manager etc.***
